# Spatiotemporal Trends of Birth Defects in North Carolina, 2003-2015

**DOI:** 10.1101/2024.08.12.24311873

**Authors:** Haidong Lu, Andrew F. Olshan, Marc L. Serre, Kurtis M. Anthony, Rebecca C. Fry, Nina E. Forestieri, Alexander P. Keil

**Affiliations:** Department of Internal Medicine, Yale School of Medicine, CT, USA; Department of Epidemiology, Gillings School of Global Public Health, University of North Carolina at Chapel Hill, Chapel Hill, NC, USA; Department of Environmental Sciences and Engineering, Gillings School of Global Public Health, University of North Carolina at Chapel Hill, Chapel Hill, NC, USA; Institute for Environmental Health Solutions, Gillings School of Global Public Health, University of North Carolina at Chapel Hill, Chapel Hill, NC, USA; Curriculum in Toxicology and Environmental Medicine, University of North Carolina at Chapel Hill, Chapel Hill, NC, USA; Birth Defects Monitoring Program, State Center for Health Statistics, North Carolina Department of Health and Human Services, Raleigh, NC, USA

**Keywords:** birth defects, Bayesian disease mapping, spatiotemporal analysis

## Abstract

Birth defects are a leading cause of infant mortality in the United States, but little is known about causes of many types of birth defects. Spatiotemporal disease mapping to identify high-prevalence areas, is a potential strategy to narrow the search for potential environmental and other causes that aggregate over space and time. We described the spatial and temporal trends of the prevalence of birth defects in North Carolina during 2003-2015, using data on live births obtained from the North Carolina Birth Defects Monitoring Program. By employing a Bayesian space-time Poisson model, we estimated spatial and temporal trends of non-chromosomal and chromosomal birth defects. During 2003-2015, 52,524 (3.3%) of 1,598,807 live births had at least one recorded birth defect. The prevalence of non-chromosomal birth defects decreased from 3.8% in 2003 to 2.9% in 2015. Spatial modeling suggested a large geographic variation in non-chromosomal birth defects at census-tract level, with the highest prevalence in south-eastern North Carolina. The strong spatial heterogeneity revealed in this work allowed to identify geographic areas with higher prevalence of non-chromosomal birth defects in North Carolina. This variation will help inform future research focused on epidemiologic studies of birth defects to identify etiologic factors.

---

Birth defects are a leading cause of infant mortality in the US.^1,2^ In North Carolina, about 3% of all births are affected by birth defects each year.^3^ In spite of the substantial health impact, with a few exceptions, little is known about modifiable causes or prevention of birth defects. Over 60% of birth defect cases have no known cause.^4^ Some factors such as chemical exposures, radiation, and medications have been associated with birth defects, leaving open the possibility that an important proportion of birth defects may be attributable to environmental causes.^5,6^ Environmental exposures to persons often occur due to emission by fixed or mobile sources, thus leading to correlated exposures of individuals who are in spatiotemporal proximity. Synthesizing spatial information and exploring spatiotemporal patterns of the occurrences of birth defects may help to identify high-risk areas and populations and narrow the search for potential environmental and other spatially situated causes.

Disease mapping, a visual representation of disease outcomes across geographic areas, has long been undertaken to facilitate description and investigation of disease outcomes and to address disease priorities. Disease mapping can also provide additional insights in highlighting high-risk populations, identifying modifiable causes of diseases, and explaining and predicting disease patterns. One barrier to progress in describing the spatial distribution of birth defect occurrence is disease rarity, especially within small areas (e.g., census tract), which leads to large uncertainty in area estimates of prevalence. Bayesian spatiotemporal modeling, which has become increasingly popular in public health research^7^ can reduce this concern under the assumption that areas and times in close proximity will have prevalence more similar to each other than to more distal areas and times. This technique reduces estimation uncertainty in a given area/time by borrowing information from neighboring areas and adjacent times, which can improve prevalence estimates of rare diseases.^8^

In this study, we applied Bayesian disease mapping techniques to analyze data from the North Carolina Birth Defects Monitoring Program which included birth defects diagnosed to North Carolina resident live births between 2003 and 2015. Our goals were to: (1) describe broad spatial and temporal trends in the prevalence of birth defects in North Carolina, and (2) assess deviations from the state-wide spatiotemporal trends in prevalence to highlight local space-time regions of concern. This descriptive analysis will help better understand existing spatiotemporal patterns as well as inform future investigations by identifying high-risk populations and priority regions in the search for environmental causes of birth defects.

## METHODS

### Study Population and Data

Data on liveborn infants with birth defects were obtained from the North Carolina Birth Defects Monitoring Program (NCBDMP). The NCBDMP is an active, statewide, population-based surveillance system operated by the State Center for Health Statistics that collects information about all medically diagnosed birth defect cases among North Carolina resident infants. Birth defect cases were identified through systematic review and abstraction of medical records by trained NCBDMP field staff. Diagnoses were confirmed by the supporting documentation in medical records (e.g., medical imaging, physical exams, autopsy reports). During the same years, birth certificate records were used to identify all live births in North Carolina. The affected and unaffected births serve as the base population of pregnancies from which affected fetuses are assumed to arise. Each record included demographic information such as maternal age at delivery and education, and infant sex, race, birth weight, multiplicity (singleton vs. other), delivery type (vaginal vs. cesarean) and gestational age at delivery. GPS-based latitude and longitude of maternal residence at delivery was recorded for all births.

In the present study, we included data on all North Carolina resident births between 2003 and 2015. The latitude and longitude coordinates for each birth were then matched to census tracts using the R tigris package.^9^ For each census tract, birth defect cases and unaffected births were aggregated to annual counts. Sixteen of 2,195 census tracts (0.7%) with zero births across 2003 to 2015 were excluded from analysis. For subsequent modeling purposes, we created an adjacency matrix, which characterizes all bordering census tracts for each census tract in North Carolina, using the R spdep package.^10^ This study was approved by the University of North Carolina at Chapel Hill Institutional Review Board under a waiver of informed consent.

### Outcomes

The primary outcome was diagnosis of any non-chromosomal birth defect. In addition, based on previous work into associations between birth defects and exposures from well water in North Carolina^6^, several individual major non-chromosomal birth defects were evaluated: 1) Anotia and microtia; 2) Conotruncal heart defects including common truncus, tetralogy of Fallot, and transposition of the great arteries; 3) Atrioventricular septal defects and endocardial cushion defects; 4) Cleft lip with or without cleft palate; 5) Cleft palate; 6) Hypospadias; 7) Gastroschisis. CDC/BPA codes for each defect are given in S1 Appendix Table 1. The prevalence of overall birth defects and chromosomal birth defects was also examined.

### Target parameters

The current analysis focuses on description, rather than causal inference, so we seek to estimate the crude (i.e., unadjusted for covariates) prevalence of non-chromosomal birth defects within each census tract-year in North Carolina. The crude prevalence is calculated by taking the number of non-chromosomal birth defects and dividing by the total number of live births. We use this crude prevalence as input into our spatiotemporal mapping scheme to estimate the annual prevalence of non-chromosomal birth defects among births with the potential to be affected and recorded by NCBMDP. This approach estimates a hypothetical “underlying” prevalence of non-chromosomal birth defects from which our data are only a single realization. The target parameter we wish to estimate is the prevalence ratio which contrasts the prevalence in a specific area and time with the average prevalence across the entire study period. Thus, a prevalence ratio > 1.0 for a given census-tract-year indicates higher prevalence than the North Carolina average over the study period. This approach can be considered an approximation of a fetuses-at-risk approach^11^ (see S2 Appendix), where we deviate from such an approach by missing information on fetal losses, and timing for each birth is defined by date of delivery rather than date of conception. Estimated crude prevalence ratios will be approximately unbiased if the proportion of fetal losses to total pregnancies is approximately constant over the study area and period.

### Statistical Analyses

We estimated annual prevalence of non-chromosomal birth defects by census tract in North Carolina using a Bayesian space-time model that is widely used in spatial epidemiology.^7,12^ We opted for this approach because birth defects are rare, and we would thus expect crude prevalence estimates within a given census tract and year to be unstable, or highly variable. With such highly variable prevalence estimates, it may be difficult to intuit spatiotemporal patterns, if they exist. Our Bayesian approach overcomes this instability by using carefully constructed priors that allow partial pooling of information across adjacent census tracts within a given calendar year, as well as by partial pooling of information across time within a given census tract. Thus, the approach assumes that the underlying prevalence of non-chromosomal birth defects varies smoothly over adjacent census tracts and years. Our general approach is to do this information borrowing without imposing strong modeling assumptions for spatial or temporal trends, which could potentially obscure important patterns.

Our modeling approach can be expressed as a multi-level model.^13^ For each non-chromosomal birth defect considered, we modelled the number of affected births *y_it_* in census tract i during year t as conditionally independent and identically Poisson distributed variables with mean given by *λ_it_*,

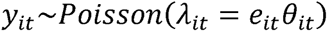

Where the mean *λ_it_* consists of two components, *e_it_* representing expected counts of non-chromosomal birth defects (described below) in the *i*th census tract during year *t*, and *θ_it_* representing the prevalence ratio for the ith census tract during year t. Then, the natural logarithm of e_it_ was modelled as

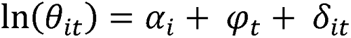

Where *α_i_* is census tract level spatial main-effect, *ψ_t_* is a temporal main-effect, and *δ_it_* is an interaction term between space (census tract level) and time. We computed the expected counts (e_it_) as the product of the number of live births in the *i*th census tract during year t and the average prevalence across the entire study period in North Carolina. Thus, the expected count estimates the number of non-chromosomal birth defects in a given census tract-year, had that census tract-year been subject to the same average prevalence as all of North Carolina from 2003 – 2015. This construction implies that *θ_it_* estimates a prevalence ratio comparing a census-tract-year prevalence to the average prevalence in North Carolina over the study period, such that values > 1 imply prevalence higher than the state average that can be used to locate potentially high-risk groups.

The spatial, temporal, and spatiotemporal interaction terms are parameterized to provide structure to the prevalence estimates without making strong modeling assumptions that might otherwise smooth over key spatial or temporal trends. The spatial term a_i_ is a random effect that follows the conditional autoregressive model proposed by Besag, York and Mollie.^14^ This random effect can be further decomposed into two components, an intrinsic conditional autoregressive term that smooths each census tract estimate by forming a weighted average with all adjacent census tracts, plus a spatially unstructured component that models independent location-specific error and is assumed to be independently, identically, and normally distributed across census tracts. The temporal trend *ψ_t_*, is modeled by the sum of two components, a first-order random walk-correlated time component (which is conceptualized as a prior in which the temporal term in year t is given a normal prior centered on the value of the temporal term in year *t*-1), and a temporally unstructured component that models independent year-specific error and is independently, identically, and normally distributed across years. The space-time interaction term *δ_it_*, is modelled as an independent noise term for each census tract and time period, and allows for temporal trends in a given census tract to deviate from the overall trend, such that spatiotemporally local patterns can emerge by reducing the amount of smoothing done by the model. Penalized complexity (PC) priors^15,16^ were applied to the precision hyperparameters in our models. Details of model specification are described in S3 Appendix.

To estimate Bayesian model parameters, we employed integrated nested Laplace approximations (INLAs) which approximate the full posterior distribution and are a computationally efficient alternative to Markov Chain Monte Carlo (MCMC) for certain model structures (latent Gaussian models). INLA does not use iterative computation techniques like MCMC and is thus highly efficient at the cost of possible approximation error.^17^ We used the R-INLA package for model fitting.^8^ Model comparison was performed, and details can be found in S4 Appendix Table 2.

The prevalence of individual non-chromosomal birth defects, any birth defect (including non-chromosomal birth defects and chromosomal birth defects), and chromosomal birth defects in North Carolina was estimated using the same Bayesian approach.

## RESULTS

Of 1,600,409 affected and unaffected births recorded in NCBDMP during the study period 2003-2015, 758 had maternal residence outside North Carolina, and 844 had inaccurate geographic information that prevented precise geocoding. After excluding these records, a total of 1,598,807 live births were included in the analyses. Among these, 52,524 (3.3%) had at least one recorded birth defect. The prevalence of any birth defect decreased from 4.0% in 2003 to 3.2% in 2015, as shown in Table 1. The prevalence of non-chromosomal birth defects decreased from 3.8% in 2003 to 2.9% in 2015. The numbers of individual structural birth defects (i.e., anotia/microtia, conotruncal heart defects, atrioventricular septal defects and endocardial cushion defects, cleft lip, cleft palate, hypospadias, and gastroschisis) are also presented in Table 1.

**Table 1.**
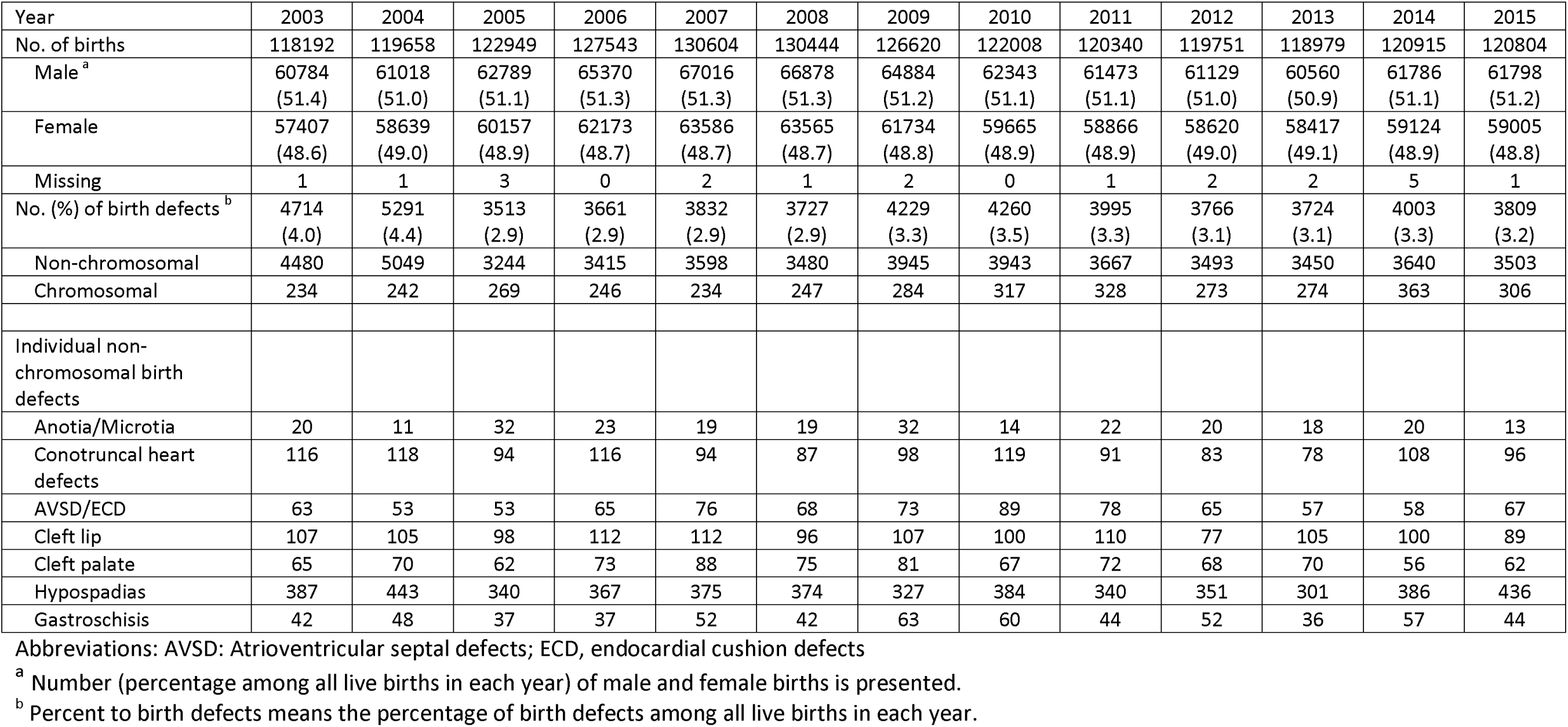
Number of births by year among 1,598,807 live births (52,524 birth defects) in North Carolina in 2003-2015.

The posterior geometric means of spatial random effect for the prevalence (“spatial prevalence ratio” – holding temporal terms constant) of any non-chromosomal birth defect are summarized in Figure 1. This map reveals a large variability of the spatial term of the model, as shown with prevalence ratio varying geographically from a low of below 0.6 to a high of about 2.0 across the state. The spatial prevalence ratio identifies areas at heightened prevalence of birth defects in North Carolina throughout the 2003-2015 period. Of note, the southeastern region of North Carolina had the highest prevalence of birth defects, though higher prevalence was also noted in the Appalachian and Northern Piedmont areas.

**Figure 1.**
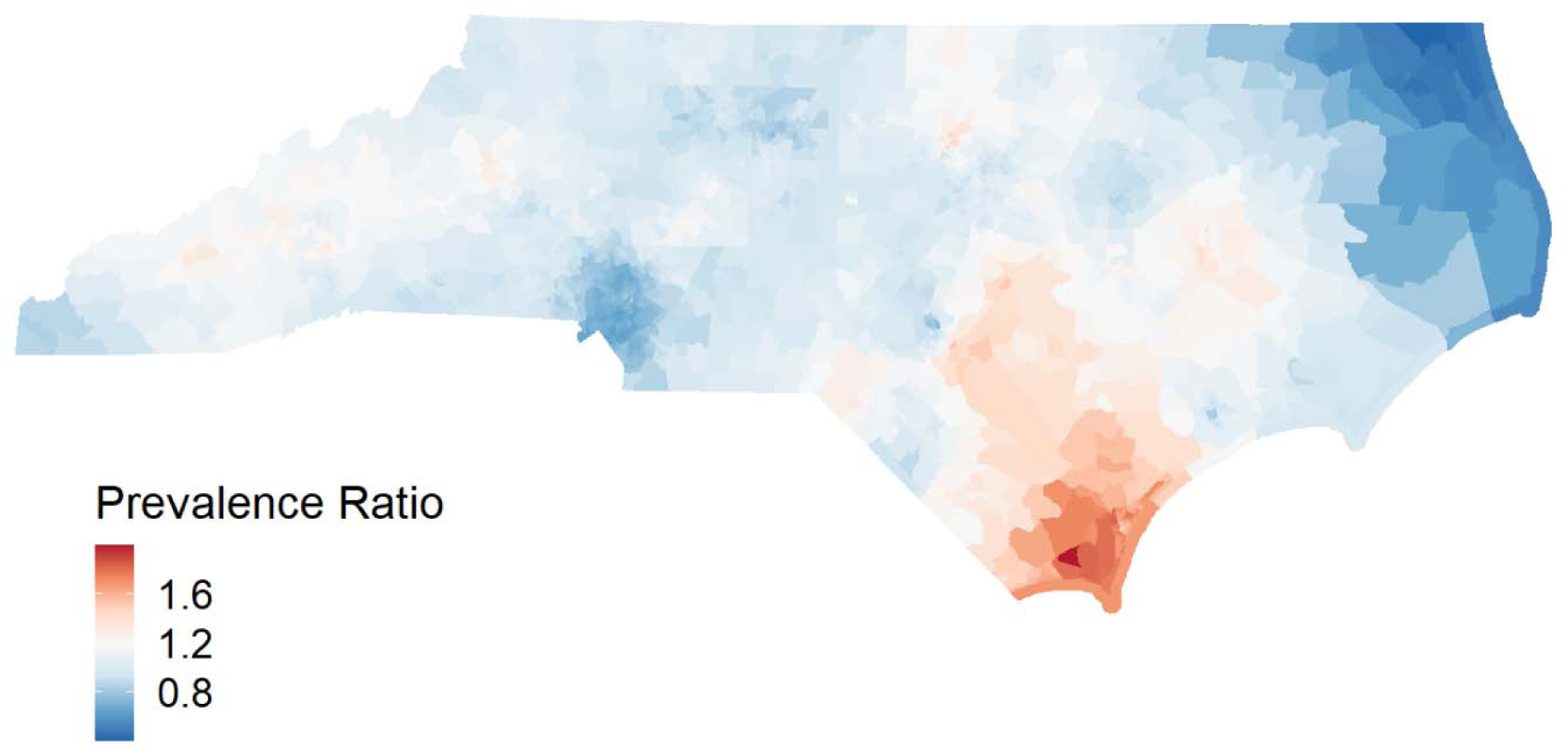
Posterior geometric mean prevalence ratio for any non-chromosomal birth defect across North Carolina, spatiotemporal model of North Carolina census tracts, 2003-2015. It represents the autoregressive spatial term.

Posterior geometric means of the temporal random effect (“temporal prevalence ratio” – holding spatial terms constant) is depicted in Figure 2. The temporal prevalence ratio was highest during the first two years (2003 and 2004), and then dropped. While there was a slight spike during 2009-2010, the overall prevalence appeared constant over time since 2005.

**Figure 2.**
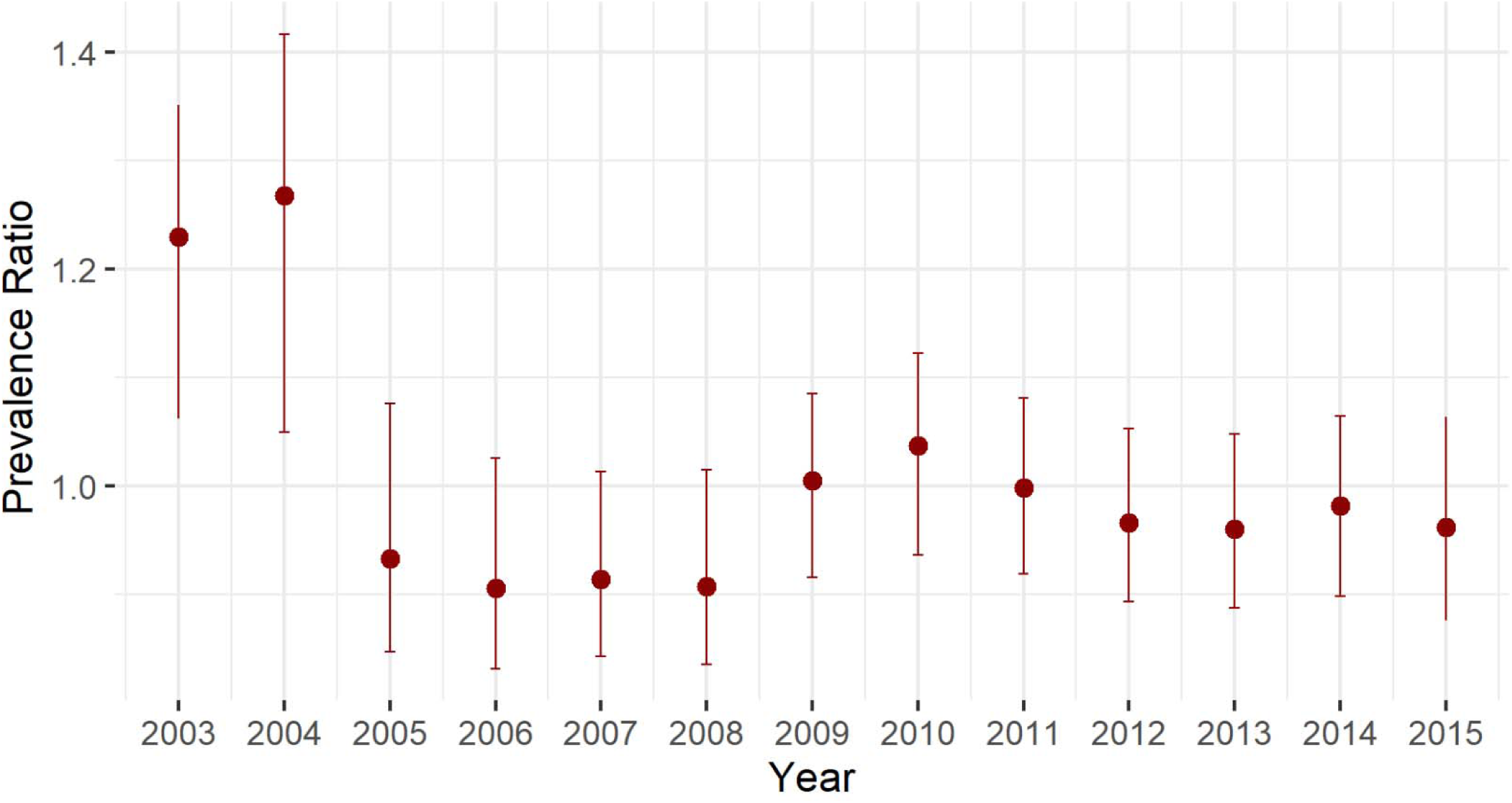
Temporal trend term of non-chromosomal birth defects, spatio-temporal model of North Carolina census tracts, 2003-2015

Posterior geometric means of the independent yearly space-time interaction term are presented for four of the study period years in Figure 3. These interactions capture local deviations from overall spatial and temporal trends. As shown in Figure 3, there are some census tracts with elevated prevalence of any birth defect in 2004. But generally, the space-time interaction term varies only from about 0.88 to 1.14 (Figure 3), which is a narrower range of variability than that of the spatial term (Figure 1). This result suggests that birth defects might be associated with factors that are purely geographical, or factors that have a stronger variation over space than time.

**Figure 3.**
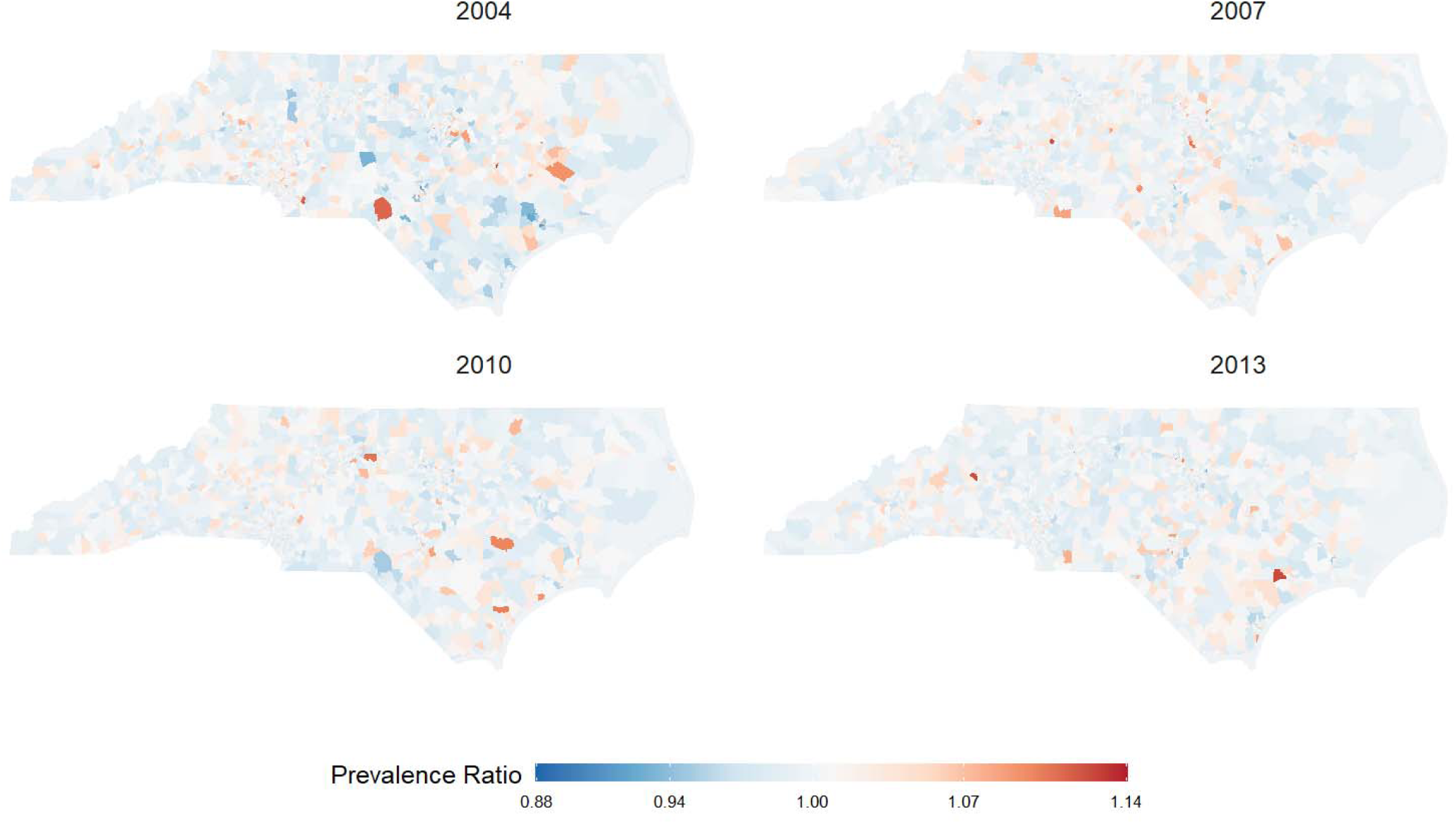
Posterior means of the independent space-time interaction term, spatio-temporal model of North Carolina census tracts, 2003-2015. Note that posterior mean log-prevalence ratios are exponentiated to represent posterior geometric mean prevalence ratios. Regions with lighter color suggest no space-time interaction and no local deviations from overall spatial and temporal trends, while regions with deeper color suggest there is space-time interaction and local shock that deviates from overall spatial and temporal trends.

The spatial and temporal patterns of individual birth defects (i.e., anotia/microtia, conotruncal heart defects, atrioventricular septal defects and endocardial cushion defects, cleft lip, cleft palate, hypospadias, gastroschisis) are depicted in S5 Appendix. Generally, the prevalence of these individual birth defects remains constant across the 2003-2015 period, suggesting that the temporal trend observed in all birth defects combined was not solely attributable to any of these specific defects. In terms of spatial heterogeneity, there was some variation in patterns for defect groups. The central and southern regions of North Carolina experienced the highest prevalence of conotruncal heart defects; the west and south parts of North Carolina had increased prevalence of cleft lip and cleft palate as well as gastroschisis; the areas with higher prevalence ratios for hypospadias were strongly concentrated in the middle (Raleigh) and southern (Wilmington) urban parts of North Carolina (see S5 Appendix).

The spatial and temporal patterns of any birth defect (including non-chromosomal birth defects and chromosomal birth defects) are depicted in S6 Appendix. The geographic distributions and temporal trends of any birth defect are similar to those of non-chromosomal birth defects. The spatial and temporal patterns of chromosomal birth defects are depicted in S7 Appendix. For chromosomal birth defects, the prevalence was higher in the middle part of North Carolina, compared with other regions. The spatial trends suggest that the prevalence of chromosomal birth defects increased after 2008.

## DISCUSSION

In the present study we examined the spatial and temporal patterns of birth defects in North Carolina during 2003-2015 using small-area Bayesian spatiotemporal models. To our knowledge, it is among the first studies to map the distributions of non-chromosomal birth defects, chromosomal birth defects, and individual birth defects over time in North Carolina. We identified some regions of North Carolina, particularly in the Southern Coastal region to have relatively high prevalence of non-chromosomal birth defects compared to the average prevalence across the state. We also found that, while the prevalence of non-chromosomal birth defects was relatively high during 2003-2004 with approximately 4% among all livebirths, the prevalence dropped down and stayed constant at about 3% in the subsequent years. Furthermore, spatial heterogeneity was also apparent for several individual birth defect groups including conotruncal heart defects, cleft lip, cleft palate, hypospadias, and gastroschisis. Although there is some commonality in relatively high prevalence of several birth defects (e.g., cleft lip, cleft palate, gastroschisis) in western and southern parts of North Carolina, the spatial patterning generally appeared to differ according to each defect.

Geographic variation in birth defects has been described in previous studies.^18–21^ We employed small-area statistical techniques and identified some areas with higher prevalence (relative to the state average) of birth defects (particularly non-chromosomal birth defects) at census-tract level. Because our analysis was descriptive in nature, we did not directly assess etiologic hypotheses. In addition, our model only included spatiotemporal terms and no terms for previously studied factors such as socioeconomic status and environmental exposures.

However, our mapping result could be used to integrate with other spatiotemporal data to inform further research on potential causes for birth defects in North Carolina. For example, a previous study of toxic metals in private wells and birth defects prevalence in North Carolina in 2003-2008, showed that the elevated manganese levels in the central part of the state were associated with a higher prevalence of conotruncal heart defects.^6^ This study was consistent with our finding that the central region of North Carolina has heightened prevalence of conotruncal heart defects. We have identified some regions that have a higher prevalence of non-chromosomal birth defects and some individual birth defects including conotruncal heart defects, cleft lip, cleft palate, hypospadias and gastroschisis, compared with other regions. Since we found that the spatial term of the birth defect model is significantly greater than the space-time interaction term, future work should focus on associations between birth defect prevalence and geographical factors, such as well water contamination that persists over long durations.

Following global trends, fewer births were recorded in the years immediately prior to the 2008 financial crash relative to the years immediately following.^22^ We estimated higher prevalence in birth defects occurring after 2008 relative to birth defects occurring 2005-2008. This pattern suggests that economic shocks may also play a role in the temporal patterns of birth defects across the state, especially if fertility patterns shift such that pregnancy becomes relatively more common among women with higher risk of affected offspring (e.g. older mothers due to delayed childbearing).^23,24^ The average maternal age at birth in our data was relatively steady between 2003 and 2009 (26.9-27.0) but rose steadily thereafter to 28.0 by 2015, which closely mirrors the patterns of chromosomal defects we observed and supports a maternal age hypothesis.

Our study had several limitations. Outcome ascertainment and classification may be a source of measurement error. Although we found that in 2003 and 2004 North Carolina experienced relatively high prevalence of non-chromosomal birth defects compared with other years, this might be due to changes in ascertainment and classification of birth defects over time. This could also apply to individual birth defects. It is likely that we captured some birth defects better than others, which can result in loss of information when identifying the regions at high prevalence of certain individual birth defects. Cleft lip and cleft palate, which are easily clinically assessed, both demonstrated spatial patterning without strong temporal trends, suggesting that measurement may underly the temporal trends observed in any birth defect. In addition, since we only adopted the information of maternal residence at delivery for geocoding, it is possible that non-differential misclassification may be introduced by the likelihood of maternal mobility during pregnancy. We also recognize that an any birth defect group that combines individual defects with different embryologic mechanisms and potential risk factors introduces etiologic heterogeneity.

Using Bayesian disease mapping techniques, our descriptive study examined the spatial and temporal patterns of birth defects in North Carolina during 2003-2015. We identified some geographic areas with increased prevalence of non-chromosomal birth defects and some individual birth defect groups at census tract level. The etiology of birth defects is multifactorial, and the causes for most defects remain unknown. Given the potential geographic variation in toxic environmental contaminants in North Carolina that are likely tied to the birth defects^6^, further studies are warranted to explore the potential environmental causes (e.g., well water contamination) for each type of birth defects.

## Data Availability

Data are available on request to the authors

## ACKNOWLEDGMENTS

This work was supported in part by the National Institute of Environmental Health Sciences (grant R01 ES029531), and a cooperative agreement from the Centers for Disease Control and Prevention (CDC; U50CCU422096) to the North Carolina Center for Birth Defects Research and Prevention, and through cooperative agreements under PA 96043, PA 02081, and FOA DD09-001 from the CDC to other Centers for Birth Defects Research and Prevention participating in the National Birth Defects Prevention Study. We are also thankful for the support from the North Carolina Center for Birth Defects Research and Prevention.

## APPENDIX

## S1 Appendix

**Appendix Table 1:** CDC/BPA codes for individual non-chromosomal birth defects.

|  |
| --- |
| Anotia/microtia: 741.01, 741.21 |
| Conotruncal heart defects |
| Common truncus: 745.00 |
| TOF: 745.20-745.21, 747.31 |
| TGA: 745.10-745.12, 745.18-745.19 |
| AVSD/endocardial cushion defects: 745.60-745.69, 745.487 |
| Cleft lip: 749.10-749.19 |
| Cleft palate: 749.00-749.09 |
| Hypospadias: 752.60-752.62 (excluding 752.61 and 752.621) |
| Gastroschisis: 756.71 |

## S2 Appendix Approximation to fetuses-at-risk approach

The crude census-tract-year specific prevalence ratios estimated in the study can be expressed as the quantity 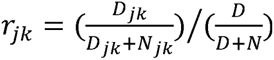, where *D_jk_* and *N_jk_* are the (observed) counts of birth defects and unaffected births in the *j*th census tract and the *k*th year, and *D* and *N* are the counts across the entire study period and area. In a fetuses-at-risk approach, the prevalence ratio would instead 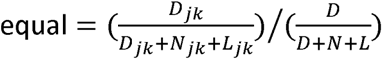, where *L_jk_*is the (unobserved) count of fetal losses *j*th census tract and the *k*th year (and, similarly *L* is the total summed over the study period and area). The prevalence of fetal losses in a given census-tract year can be expressed as 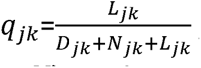, and we denote the average prevalence over the entire study period and area as *q*. Note that *L= q/1 – q)* * *(D + N)*, so that we can express the prevalence ratio as a function of observed data and the odds of fetal loss *o= q/(1 – q)*

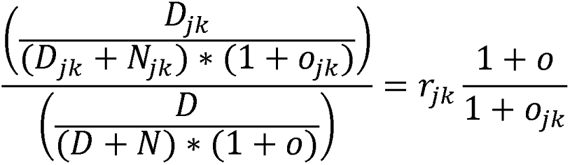

which reduces to our crude prevalence ratio in the case where the prevalence odds of fetal loss are constant across all study censustract-tract-years (*o*= *oj_k_*, which is implied by *q*= *q_jk_*, for all *j,k*). We note that, for this condition to hold, the census-tract-year specific which reduces to our crude prevalence ratio in the case where the prevalence odds of fetal loss are constant across all study census-probability of fetal loss would necessarily be inversely related to the census-tract-year specific probability of a birth defect. We loss. In this case, higher values of *r_jk_* would generally imply higher values of *o_jk_*, so that, had we been able to include fetal losses in expect it is more likely that the opposite is true and that some spatially related causes of birth defects will also be causes of fetal the data, our estimates of r_jk_ would in general be smaller than those reported in our analysis. Thus, shared causes of fetal loss and fetal death likely result in bias away from the null of census-tract-year specific prevalence ratios.

## S3 Appendix Priors on random effects in Bayesian space-time Poisson model for overall opioid-detected overdose deaths

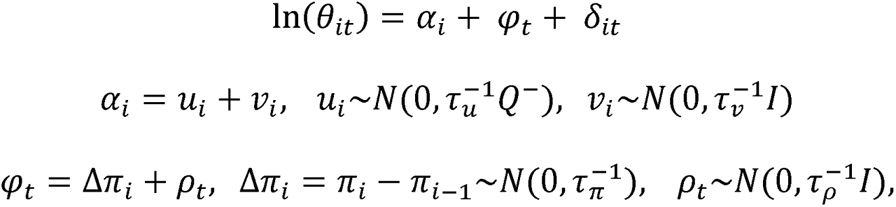

where 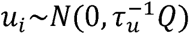 represents the spatial structured random effect and is modeled under the class of intrinsic Gaussian Markov random fields models. *Q* denotes the precision matrix (neighboring matrix), and *Q-* is the generalized inverse of the matrix *Q*. The marginal variances are 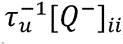 which are dependent on the matrix 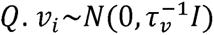 is the spatial unstructured random effect and 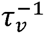 is the marginal variance. Penalized complexity (PC) priors are assigned to *ꚍ_u_* and *ꚍ_v_*. Here, we let *ꚍ_u_*, *ꚍ_v_* ∼*PC*(0.2/0.31,0.01), which corresponds to 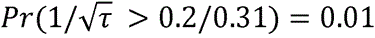.

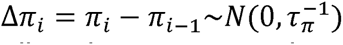 is first order random walk temporal random effect defined as a random step at each point in time (Δ*π_i_*). All random steps are independent and identically distributed. 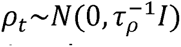 is the temporal unstructured random effect and 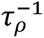 is the marginal variance. Penalized complexity (PC) priors are assigned to *ꚍ_π_* and *ꚍ_p_*. Here, we let *ꚍ_π_*, ,*ꚍ_p_* ∼*PC*(0.2/0.31,0.01), which corresponds to 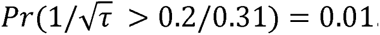.

## S4 Appendix

**Appendix Table 2:** Results of model comparison.

|  | Deviance Information Criterion (DIC) |  |  |  |  |  |  |  |  |  |
| --- | --- | --- | --- | --- | --- | --- | --- | --- | --- | --- |
|  | Defect |  |  |  |  |  |  |  |  |  |
| Model | Anotia | AVSD/E<br>CD | Cleft lip<br>w/wo<br>Cleft<br>palate | Cleft<br>palate | Conotru<br>ncal<br>heart<br>defects | Gastros<br>chisis | Hyposp<br>adias | Non-<br>chromo<br>somal | Chromo<br>somal | Any<br>defect |
| Conditional autoregressive term<br>for census tract + linear calendar<br>year + unstructured temporal<br>term for calendar year | 2872.5 | 7847.5 | 10351.1 | 7836.8 | 10212.8 | 5757.8 | 25911.7 | 85803.3 | 21303.6 | 88111.8 |
| Conditional autoregressive term<br>for census tract + first order<br>random walk term for calendar<br>year | 2871.1 | 7486.3 | 10346.7 | 7834.7 | 10213.8 | 5757.3 | 25879.7 | 85803.3 | 21302.5 | 88111.7 |
| Conditional autoregressive term<br>for census tract + first order<br>random walk term for calendar<br>year + unstructured temporal<br>term for calendar year | 2871.1 | 7486.2 | 10346.8 | 7834.8 | 10213.6 | 5757.3 | 25875.0 | 85803.2 | 21302.4 | 88112.4 |
| Conditional autoregressive term<br>for census tract + first order<br>random walk term for calendar<br>year + space-time interaction<br>term | 2872.3 | 7486.2 | 10349.4 | 7835.7 | 10211.5 | 5757.8 | 25886.9 | 85784.5 | 21301.4 | 88096.7 |
| Conditional autoregressive term<br>for census tract + first order<br>random walk term for calendar<br>year + unstructured temporal<br>term for calendar year + space-time interaction term | 2871.1 | 7486.2 | 10349.0 | 7834.7 | 10213.5 | 5757.2 | 25877.4 | 85780.2 | 21303.5 | 88094.2 |

Abbreviations: AVSD/ECD, atrioventricular septal defect/endocardial cushion defect.

## S5 Appendix: Spatial and temporal patterns of individual non-chromosomal birth defects

**Appendix Figure 5.1.**
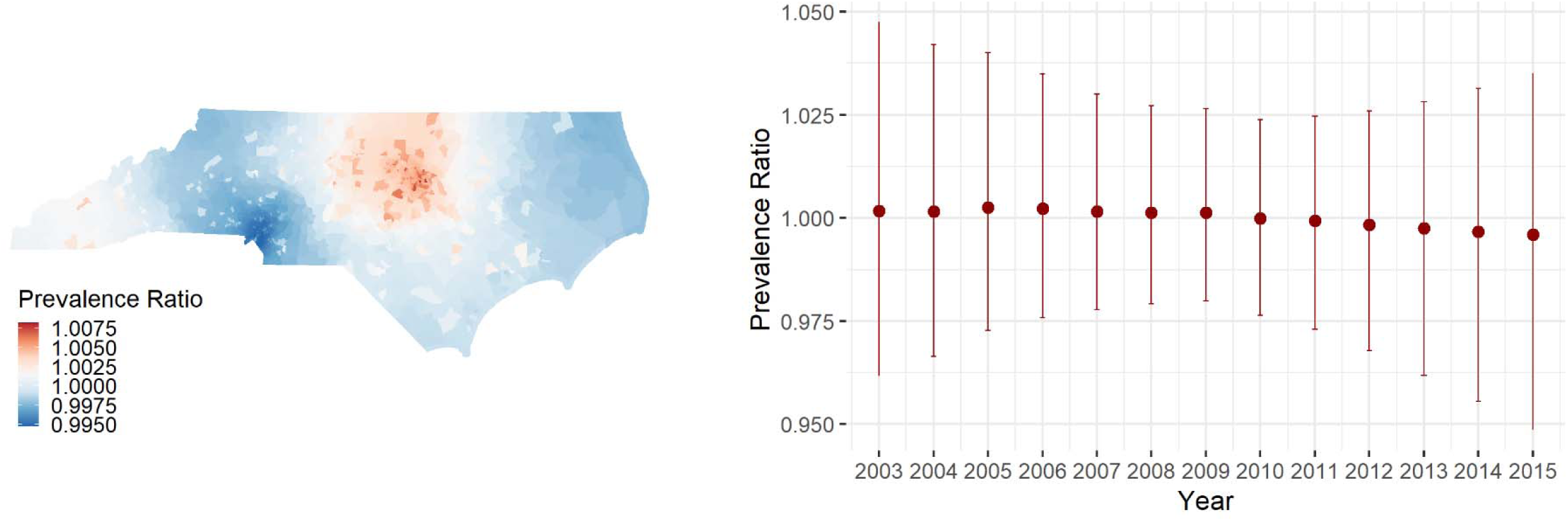
Spatial and temporal patterns of individual birth defect – anotia/microtia, North Carolina, 2003-2015

**Appendix Figure 5.2.**
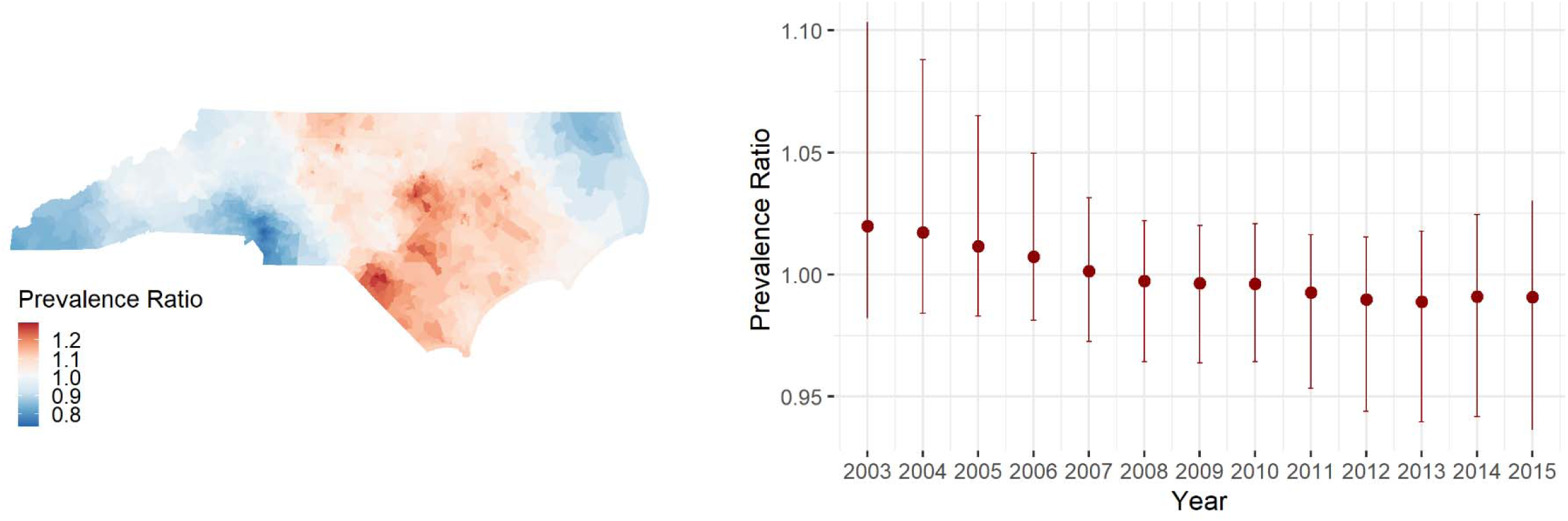
Spatial and temporal patterns of individual birth defect – conotruncal heart defects, North Carolina, 2003-2015

**Appendix Figure 5.3.**
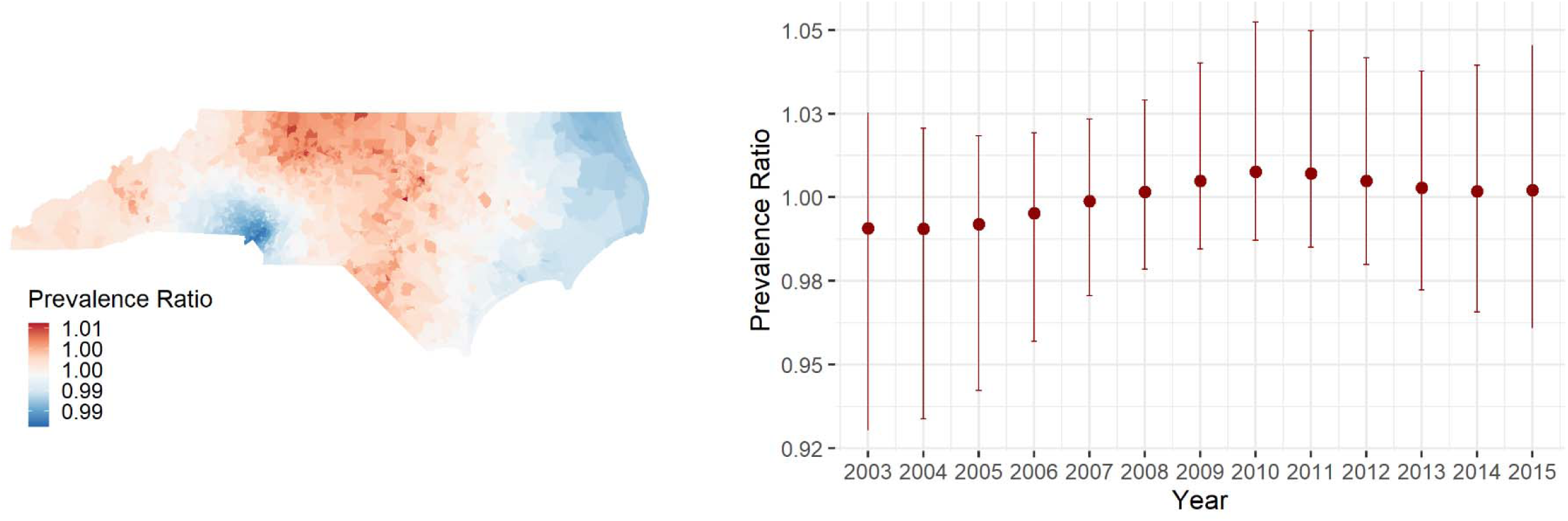
Spatial and temporal patterns of individual birth defect – atrioventricular septal defects and endocardial cushion defects, North Carolina, 2003-2015

**Appendix Figure 5.4.**
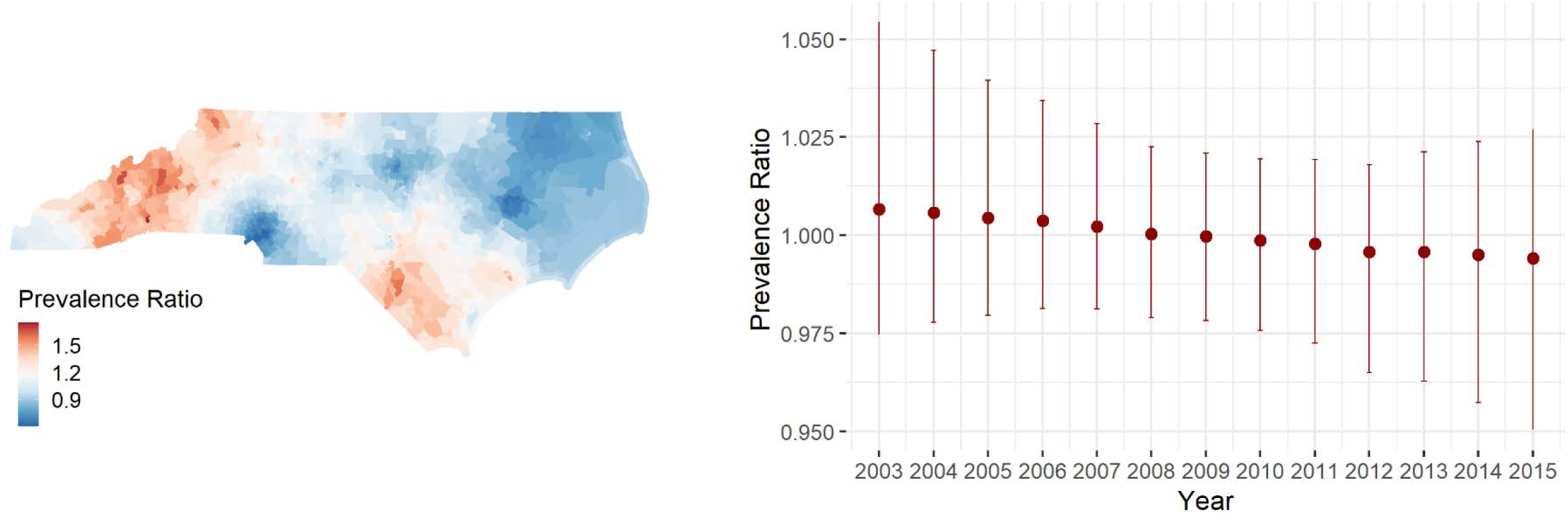
Spatial and temporal patterns of individual birth defect – cleft lip, North Carolina, 2003-2015

**Appendix Figure 5.5.**
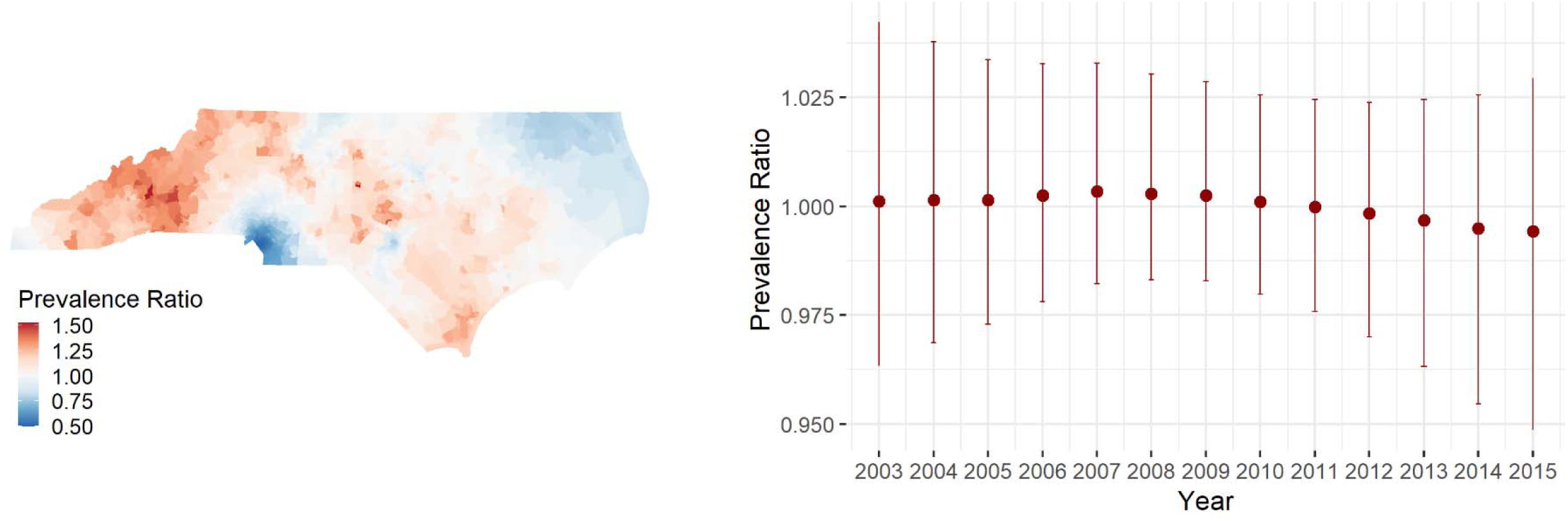
Spatial and temporal patterns of individual birth defect – cleft palate, North Carolina, 2003-2015

**Appendix Figure 5.6.**
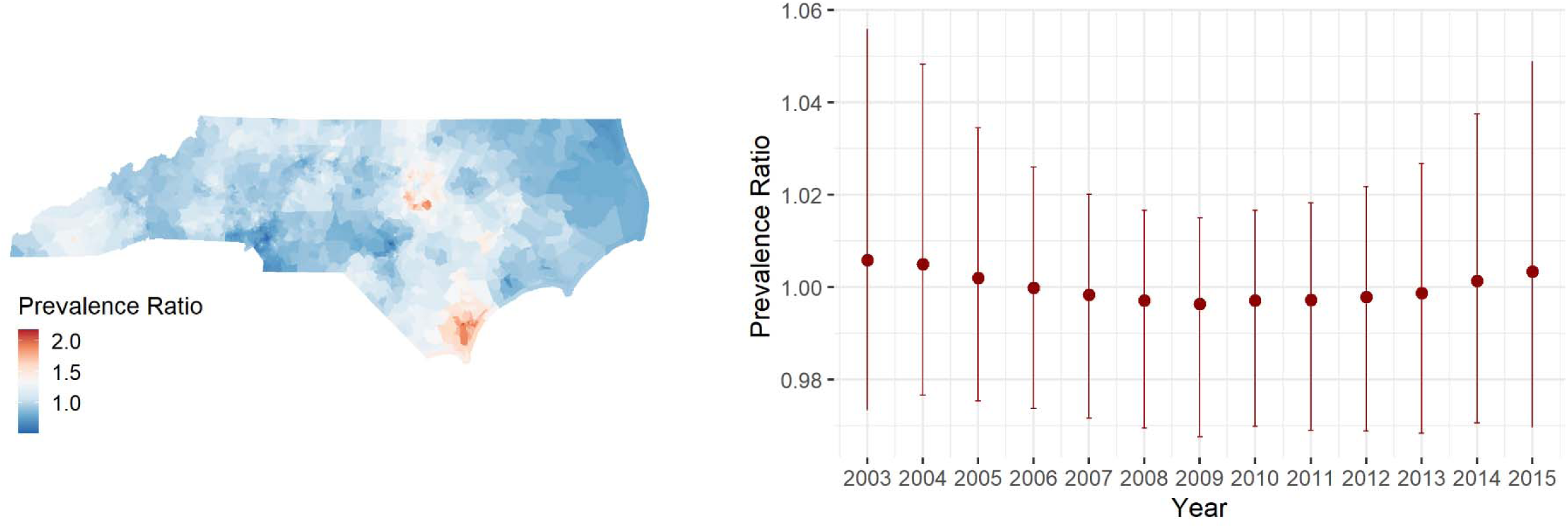
Spatial and temporal patterns of individual birth defect – hypospadias, North Carolina, 2003-2015

**Appendix Figure 5.7.**
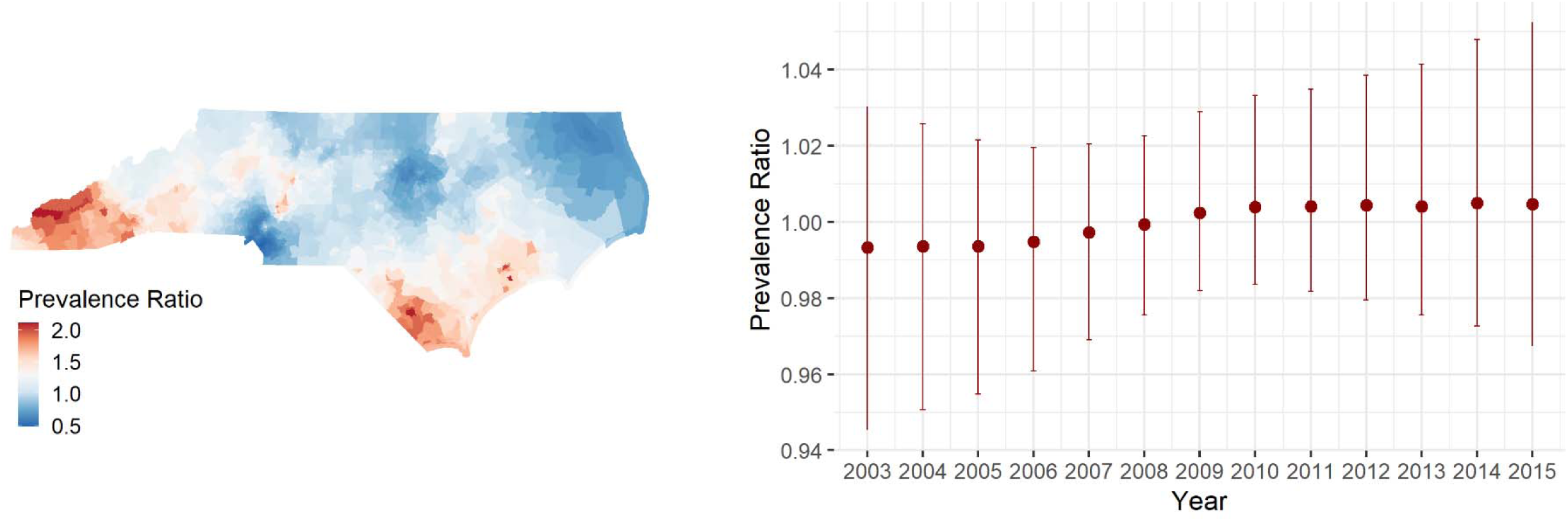
Spatial and temporal patterns of individual birth defect – gastroschisis, North Carolina, 2003-2015

## S6 Appendix: Spatial and temporal patterns of any birth defect including non-chromosomal and chromosomal birth defects

**Appendix Figure 6.1.**
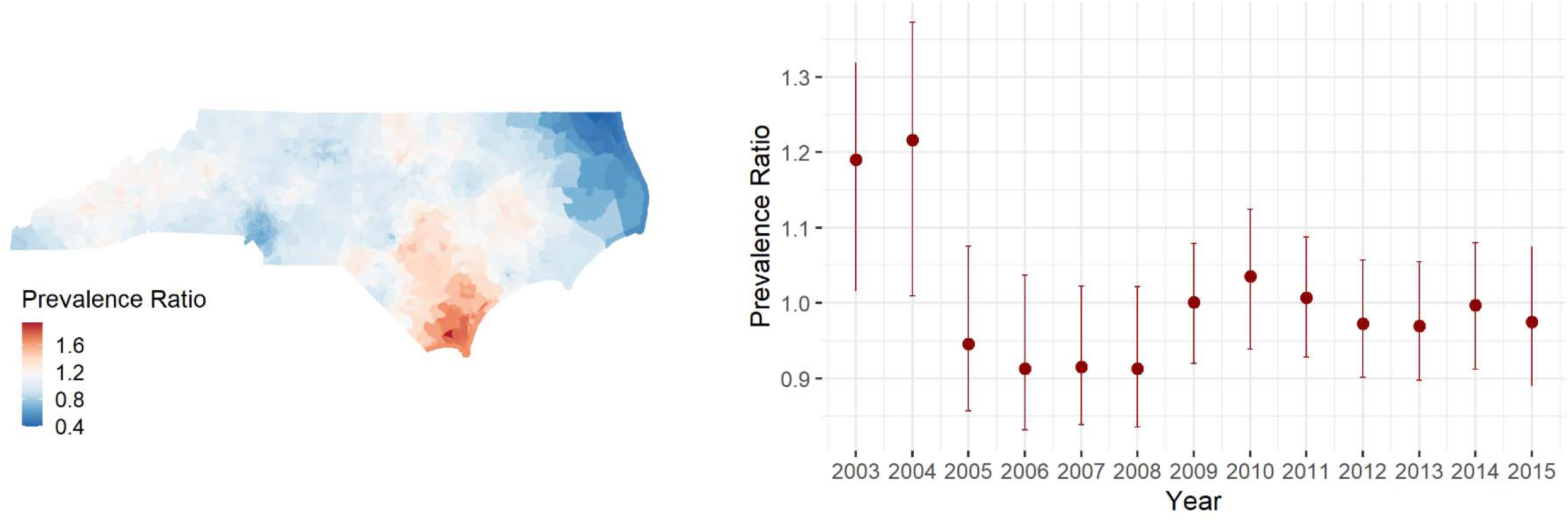
Spatial and temporal patterns of any birth defect, North Carolina, 2003-2015

## S7 Appendix: Spatial and temporal patterns of chromosomal birth defects

**Appendix Figure 7.1.**
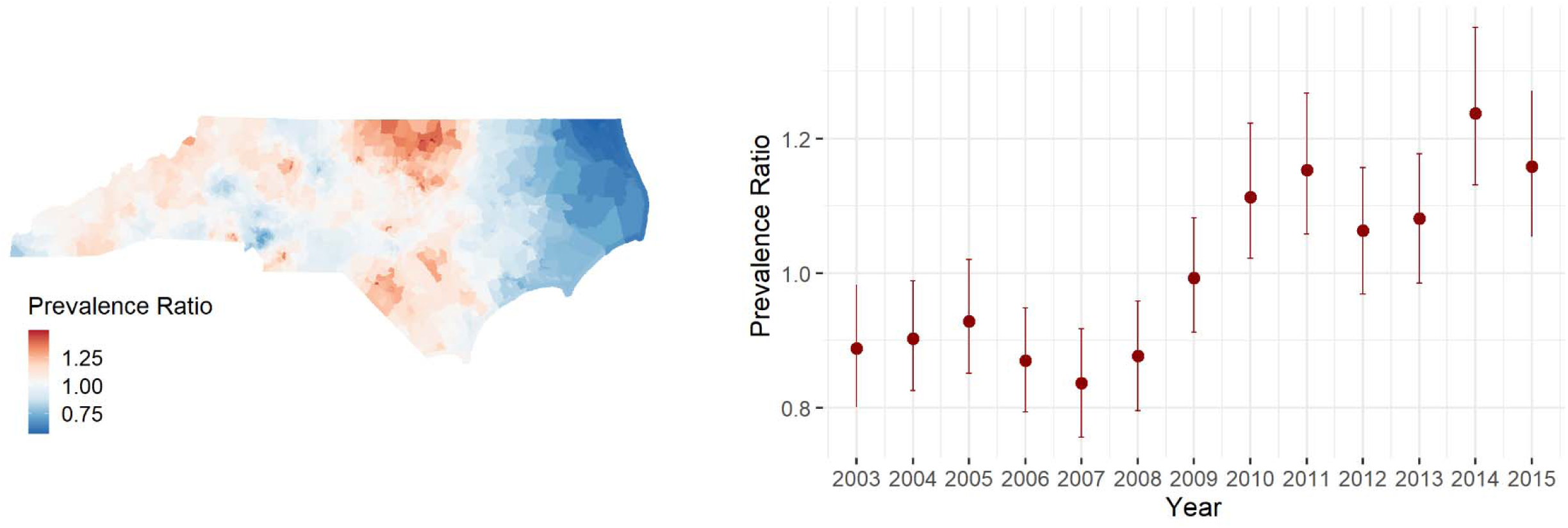
Spatial and temporal patterns of chromosomal birth defects, North Carolina, 2003-2015

